# Childhood adverse life events and skeletal muscle mitochondrial function

**DOI:** 10.1101/2023.11.07.23298177

**Authors:** Kate A. Duchowny, Theresa Mau, L. Grisell Diaz-Ramierz, Li-Yung Lui, David J. Marcinek, Frederico G. S. Toledo, Peggy M. Cawthon, Russell T. Hepple, Philip A. Kramer, Anne B. Newman, Stephen B. Kritchevsky, Steven R. Cummings, Paul M. Coen, Anthony J. A. Molina

## Abstract

Social stress experienced in childhood is associated with adverse health later in life. Mitochondrial function has been implicated as a mechanism for how stressful life events “get under the skin” to influence physical wellbeing. Using data from the Study of Muscle, Mobility and Aging (n=879, 59% women), linear models examined whether adverse childhood events (i.e., physical abuse) were associated with two measures of skeletal muscle mitochondrial energetics in older adults: (1) maximal adenosine triphosphate production (ATP_max_) and (2) maximal state 3 respiration (Max OXPHOS). Forty-five percent of the sample reported experiencing 1+ adverse childhood event. After adjustment, each additional event was associated with -0.07 SD (95% CI= - 0.12, -0.01) lower ATP_max_. No association was observed with Max OXPHOS. Adverse childhood events are associated with lower ATP production in later life. Findings indicate that mitochondrial function may be a mechanism in understanding how early social stress influences health in later life.

## INTRODUCTION

Stressful life events experienced early in the life course have been repeatedly shown to be associated with worse health later in life.^1–5^ Yet, the specific mechanism underlying this relationship remains poorly understood. Mitochondrial function (“bioenergetics”), defined by changes in the capacity to generate cellular energy in the form of adenosine triphosphate (ATP), becomes impaired with age.^6^ Recently, mitochondrial function has been implicated as a possible mechanism for understanding how stressful life events “get under the skin” to influence health and physical wellbeing.^7–12^

Mitochondria are capable of sensing and integrating social stress on a cellular level.^7^ In response to psychological stress, mitochondria undergo dynamic changes in their structure, leading to alterations in function.^7^ One potential pathway by which early childhood stress may lead to changes in mitochondrial functional capacity may involve an impaired glucocorticoid response.^4^ Persistent oxidative stress that arises from chronic, low-grade inflammation due to a blunted glucocorticoid response has been shown to be associated with mitochondrial dysfunction.^11^ Specifically, excessive glucocorticoid levels increase calcium buffering capacity, mitochondrial membrane potential, and apoptotic signaling resistance.^11^

A small number of human studies suggest that stressful social experiences may be associated with changes in mitochondrial function. For example, chronic stress due to caregiving for a family member with a chronic health condition was associated with lower mitochondrial respiratory capacity among women.^13^ Childhood maltreatment was found to be associated with changes in mitochondrial cellular respiration related to ATP production.^14^ New mothers who had a history of childhood maltreatment had alterations in ATP production and women with a more severe history of maltreatment had a worse mitochondrial bioenergetic profile.^15^

While these studies provide initial evidence that mitochondrial function may serve as a salient mechanism linking life events in childhood to poor health, two critical gaps remain. First, the majority of studies conducted have utilized measures of mitochondrial function obtained from whole blood.^9,13,14,16^ Thus, the relationship between adverse childhood events and tissue-specific measures of mitochondrial energetics obtained from skeletal muscle biopsies, the current gold-standard, is not known. Second, prior work in this area has been hampered by small sample sizes,^14^ were exclusively conducted in women^13,15^ or had minimal or no covariate adjustment. ^13–15^

In this study, we leverage data from the Study of Muscle, Mobility, and Aging (SOMMA) to examine how stressful life events in childhood may be associated with skeletal muscle mitochondrial function in later life. We extend prior studies by including in-vivo and in-vitro measures of mitochondrial energetics obtained from ^31^P MRS and muscle biopsies, in a large, well characterized sample of older adults. We hypothesized that individuals who have experienced greater adverse life events in childhood would have worse mitochondrial bioenergetic profiles in muscle (lower respiratory capacity and lower ATP_max_) in older age.

## METHODS

### Data and Sample Population

The Study of Muscle, Mobility and Aging (SOMMA) is a longitudinal, multicenter study of 879 older adults recruited between April 2019 and December 2021 from the University of Pittsburgh and Wake Forest University School of Medicine. Eligibility included age of 70 years or older, BMI less than or equal to 40 kg/m^2^, dementia free, no contraindication to a muscle tissue biopsy and magnetic resonance spectroscopy, and the ability to walk 400 meters. Participants who appeared as they might not be able to complete the 400m walk at the in-person screening visit completed a short distance walk (4 meters) to ensure their walking speed as >=0.6m/s. Participants who reported active malignancy or advanced chronic disease and/or reported that they were unable to walk ¼ mile or climb a flight of stairs were excluded. Full study details have been described elsewhere.^17^ Among the 754 participants who had data on childhood adverse life events, 698 and 633 participants had complete ATP_max_ and maximal oxidative phosphorylation (Max OXPHOS) data, respectively (see Supplementary Figure 1). All SOMMA participants provided written informed consent, and the study was approved by the WIRB-Copernicus Group (WCG IRB) (20180764).

### Measures

#### Primary Exposure: Childhood adverse life events (cALEs)

As part of the general baseline questionnaire, all study participants were asked to complete a series of questions based on a modified version of the Adverse Childhood Events scale that assesses the number of adverse life events experienced during childhood (“While you were growing up (under 18) did you experience any of the following?”).^18^ Response items included ‘Yes’, ‘No’, ‘Don’t know, Prefer not to answer’. Items ranged from physical abuse (“Did a parent or other adult in household ever slap, hit, beat, kick, or physically hurt you in any way?) to emotional neglect (“Did you often feel that no one in your family loved you or thought you were important or special?”). A total of 6 items were summed to create a continuous measure (0-6). All items are reported in the Supplement.

#### Primary Outcome Measures

Two measures of skeletal muscle mitochondrial function were utilized in this study.

1. Maximal oxidative phosphorylation (Max OXPHOS) supported by complex I and II. To obtain the in vitro Max OXPHOS measure, high resolution respirometry was performed on permeabilized fresh muscle fiber (PMF) bundles. Participants underwent a percutaneous skeletal muscle biopsy in the presence of local anesthesia, and samples were processed as previously described.^19^ Briefly, a Bergstrom canula with suction was used to collect the specimen from the middle region of the musculus vastus lateralis. The specimen was cleaned, and dissected to obtain a small bundle of myofibers that were placed into ice-cold BIOPS media (10mM Ca–EGTA buffer, 0.1M free calcium, 20mM imidazole, 20mM taurine, 50mM potassium 2-[N-morpholino]-ethanesulfonic acid, 0.5mM dithiothreitol, 6.56mM MgCl_2_, 5.77mM ATP, and 15mM phospho-creatine [PCr], pH 7.1). Myofiber bundles weighing approximately 2-3 mg were teased apart for chemical permeabilization with saponin. After washing, the PMF bundles were assayed with a standardized substrate uncoupler inhibitor titration (SUIT) protocol in duplicate to assess mitochondrial respiration as described previously.^19^ Very briefly, an Oxygraph 2K instrument (Oroboros Inc., Innsbruck, Austria) was used to measure the respiration of the PMF bundles. Specifically, for the measurement of maximal OXPHOS supported by complex I and II (also referred to as Max OXPHOS or State 3 respiration), the following substrates were added: pyruvate (5mM), malate (2mM), glutamate (10mM), succinate (10mM), and ADP (4.2mM). Data was analyzed using Datlab 7.4 software and steady-state O_2_ flux was normalized to fiber bundle wet weight.
2. Maximal ATP production (ATP_max_) was assessed in vivo using 31-Phosphorous magnetic resonance spectroscopy (^31^P MRS) as previously described. Briefly, participants lie in a supine position to perform the first bout of repeated isometric knee extension (30 sec) against the resistance of an ankle strap. A second bout was adjusted for the length of time of the kicking (18-36 sec) to achieve adequate phosphocreatine (PCr) breakdown while maintaining pH<6.8 to avoid acidic conditions inhibiting mitochondrial ATP production.^20^ Data was analyzed in jMRUI v6.0 using a standard value of 24.5mM for resting PCr. The post-exercise recovery PCr levels were used to calculate rates of mitochondrial ATP resynthesis as previously described to obtain ATP_max_ .^21,22^

### Covariates

The following covariates were considered as potential confounders or mediators and entered into the model: Age, in years; gender, study site, Pittsburgh vs. Wake Forest for ATP_max_ models, or technician group for Max OXPHOS models; education, a categorical variable that included less than high school/high school, some college, college and post-college; parental education, coded the same way as respondent education and only; educational attainment, highest number of years of schooling completed from either parent was used, a proxy measure of childhood socioeconomic position;^23,24^ body mass index (BMI) defined as weight divided by height in kg/m^2^; number of depressive symptoms, a continuous variable as determined as the Center for Epidemiologic Studies Depression Scale (CES-D);^25^ smoking status, a categorical variable defined as “Never”, “Past” and “Current”; physical activity, as assessed by the Community Healthy Activities Model Program for Seniors (CHAMPS) questionnaire, a continuous variable based on self-reported physical activity used to calculate total energy expenditure per week (Cal/week);^26^ and, total number of chronic conditions, a summary multimorbidity index based on the Rochester Epidemiology Project, scored 0-12.^27^

### Analytic approach

Outcome variables were normally distributed and standardized to facilitate comparisons. We employed linear regression models that examined the association between continuous childhood ALE and both Max OXPHOS and ATP_max_ in separate models in the overall sample. Given initial evidence suggesting there may be gender differences in mitochondrial bioenergetics,^19,28,29^ we also tested for statistical interaction by gender. We proceeded with the following modeling strategy: Model 1 (Base model) adjusted for age, gender (overall model), site and/or technician. Model 2 (Confounder model) adjusted for Model 1 and parental education, a hypothesized confounder that is associated with the childhood ALEs and potentially associated with both mitochondrial outcomes measures but precedes the exposure. Lastly, Model 3 (Confounder/Mediator model) adjusted for Model 2 and variables that could be hypothesized as confounders but are also downstream of the exposure and on the causal pathway, and therefore could plausibly serve as potential mediators, which included: participant’s education, smoking status, depressive symptoms, BMI, physical activity, and number of chronic conditions. We note that this final model may represent over control given the adjustment for potential mediators and results should therefore be interpreted with caution.^30^ We also fitted these three gender-stratified models. After accounting for missingness across our primary exposure and outcome variables (ATP_max_, n= 698 and OXPHOS, n= 633), we performed a complete case analysis. Missingness was between 4-5% across all models. Analyses were conducted using SAS 9.4 (Cary, NC).

## RESULTS

Of the 879 participants included in the study, 59% were women, the mean chronologic age was 76.3 years (SD= 5 years) and 86% self-reported white. While about half of the sample had completed some college or obtained a college degree (49.9%), only 32% of the respondent’s parents completed some college or college. Among the 754 participants who had complete data on cALEs, 53% reported experiencing 1 or more adverse events in childhood. Among those experiencing 3+ events, 55 individuals reporting experiencing 3 events, 29 participants, 4 events; 25 participants, 5 events; and, 8 participants reported experiencing all 6 events. Individuals who reported experiencing stressful life events in childhood, on average, reported a greater number of depressive symptoms. See Table 1.

**Table 1.**
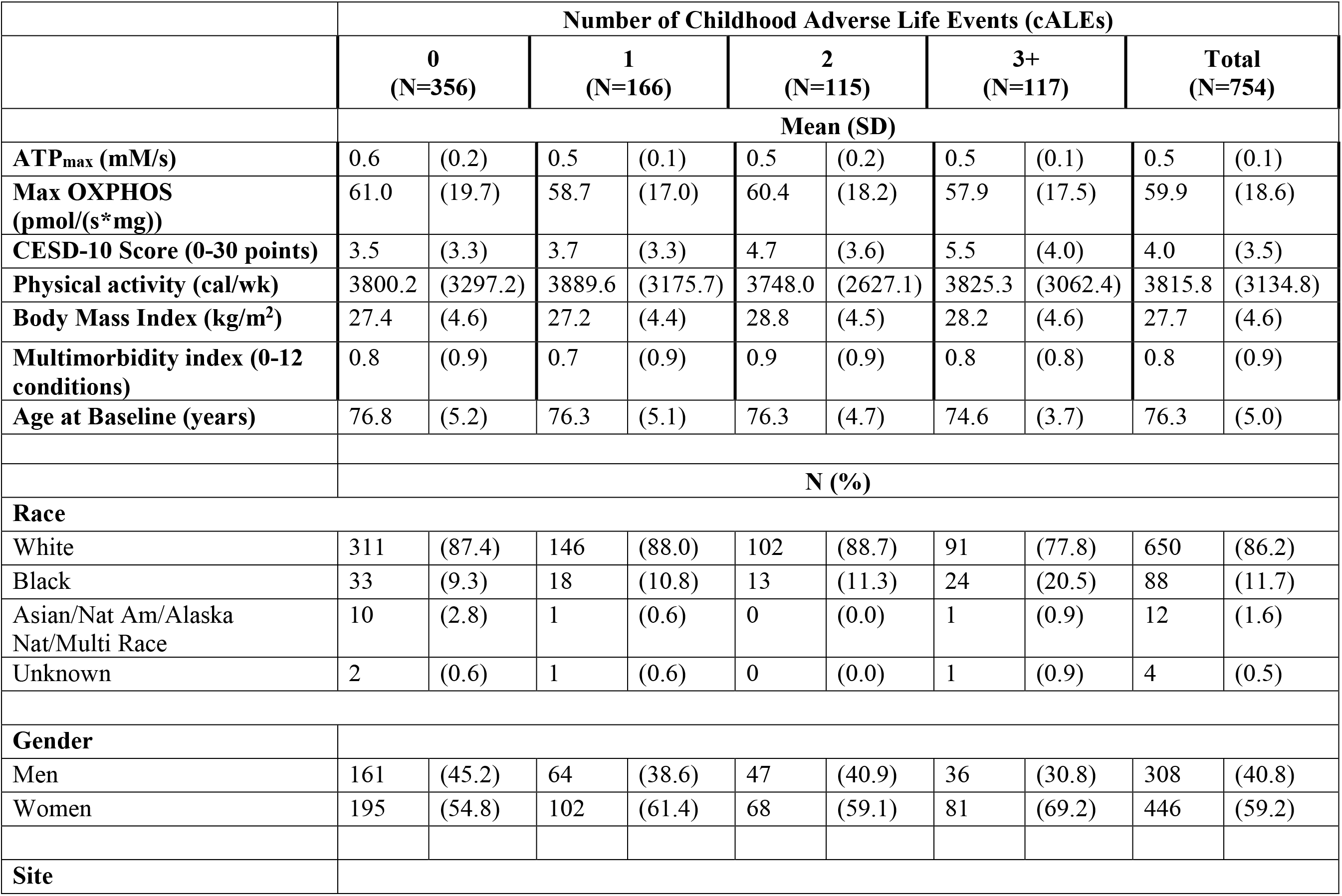
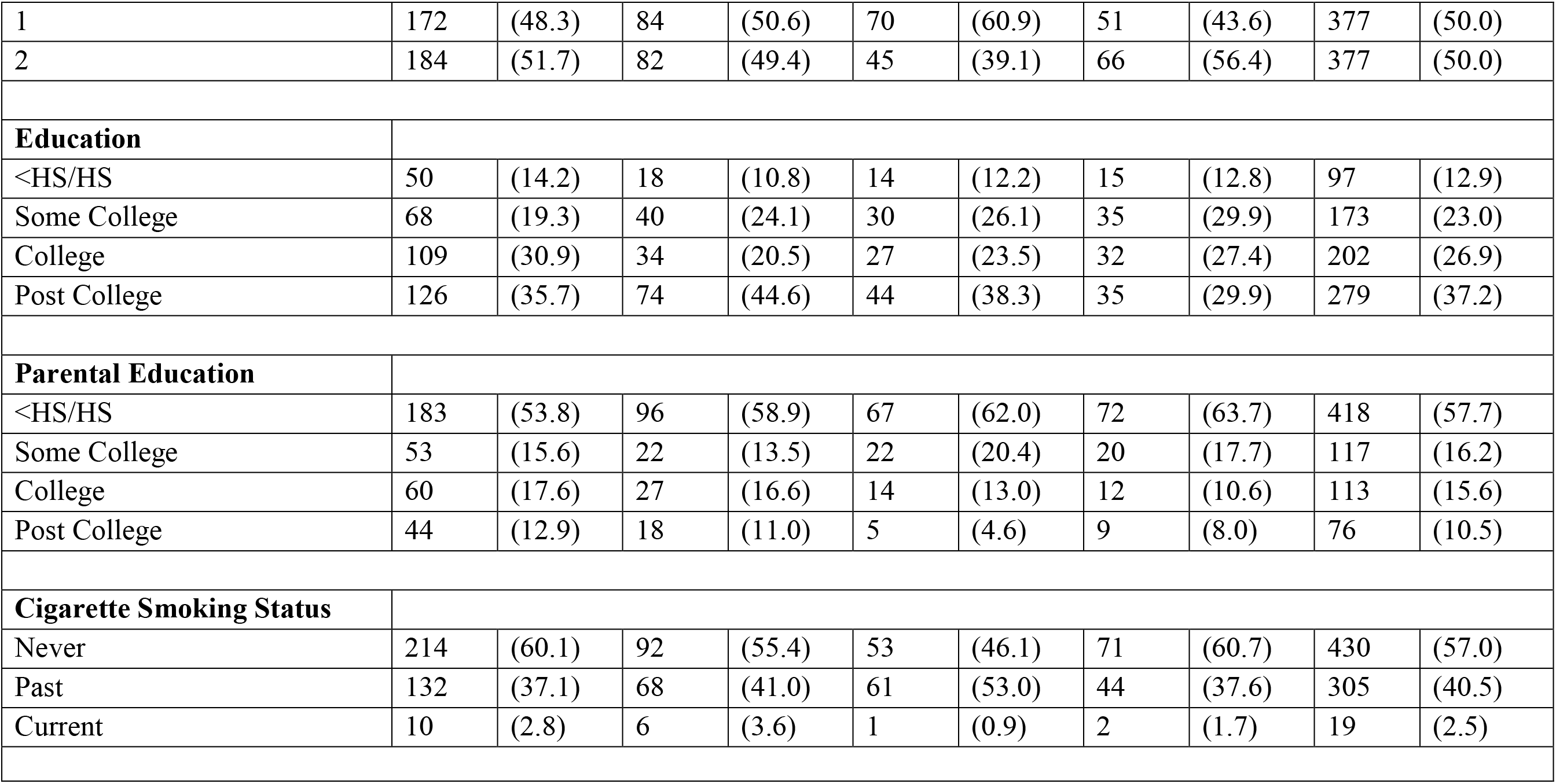
Baseline Sample Characteristics by Childhood Adverse Life Events in the SOMMA Study (n=879).

### Results for overall sample

After adjusting for age, site, and gender (Model 1), each additional childhood adverse life event was associated with a -0.10SD lower ATP_max_ (95% CI= -0.19, - 0.01). Adjusting for parental education (Model 2) and hypothesized confounder-mediators (Model 3) slightly attenuated the association such that each additional adverse event was associated with a -0.07SD lower ATP_max_ (95% CI= -0.12, -0.01). When examining effect modification by gender, the interaction term was not significant for ATP_max_ (p=0.36) nor for Max OXPHOS (p=0.78), however, given previous reports of potential sex differences in mitochondria,^28,29^ we analyzed gender stratified models (Tables 2A and 2B) and report results for men and women below. The associations with Max OXPHOS were not statistically significant across all models.

### Results for women

After adjusting for age and site (Model 1), each additional adverse life event experienced before the age of 18 was associated with a -0.04SD lower ATP_max_ (95%= -0.10, 0.02) although the confidence interval included the null value. The results, when adding parental education (Model 2) and further adjusting for hypothesized confounder-mediators (Model 3), did not change the overall association (Table 2A). While these results were not statistically significant, the effect estimates, while small, were in the anticipated direction and inversely related to cALEs.

**Table 2.** Overall linear regression results examining the relationship between childhood adverse life events and standardized ATP_Max_.

|  | **Model 1** |  | **Model 2** |  | **Model 3** |  |
| --- | --- | --- | --- | --- | --- | --- |
|  | **β** | **(95% CI)** | **β** | **(95% CI)** | **β** | **(95% CI)** |
| **Intercept** | 2.04 | [0.88, 3.20] | 1.99 | [0.80, 3.19] | 1.16 | [-0.26, 2.57] |
| **Childhood Adverse Life Events (cALEs)** | -0.10 | [-0.19, -0.01] | -0.06 | [-0.11, -0.00] | -0.07 | [-0.12, -0.01] |
| **Age (years)** | -0.02 | [-0.04, -0.01] | -0.02 | [-0.04, -0.01] | -0.01 | [-0.03, 0.00] |
| **Site<sup>a</sup>** | 0.25 | [0.10, 0.39] | 0.28 | [0.12, 0.43] | 0.30 | [0.15, 0.45] |
| **Gender<sup>b</sup>** | -0.37 | [-0.56, -0.18] | -0.32 | [-0.47, -0.16] | -0.22 | [-0.38, -0.06] |
| **Gender\*cALEs** | 0.05 | [-0.06, 0.16] |  |  |  |  |
| **Parental Education<sup>c</sup>** |  |  |  |  |  |  |
| Some college |  |  | -0.13 | [-0.35, 0.08] | -0.16 | [-0.37, 0.05] |
| College |  |  | 0.06 | [-0.16, 0.28] | -0.03 | [-0.25, 0.19] |
| Post College |  |  | 0.04 | [-0.21, 0.29] | -0.04 | [-0.30, 0.22] |
| **Body Mass Index (kg/m<sup>2</sup>)** |  |  |  |  | -0.01 | [-0.03, 0.00] |
| **CESD-10 Score (0-30 points)** |  |  |  |  | 0.02 | [-0.00, 0.05] |
| **Education<sup>c</sup>** |  |  |  |  |  |  |
| Some college |  |  |  |  | 0.14 | [-0.13, 0.41] |
| College |  |  |  |  | 0.32 | [0.04, 0.59] |
| Post College |  |  |  |  | 0.24 | [-0.02, 0.51] |
| **Physical activity (cal/wk)** |  |  |  |  | 0.00 | [0.00, 0.00] |
| **Multimorbidity index (0-12)** |  |  |  |  | -0.10 | [-0.20, -0.01] |
| **Smoking Status<sup>d</sup>** |  |  |  |  |  |  |
| Former |  |  |  |  | -0.08 | [-0.23, 0.07] |
| Current |  |  |  |  | -0.52 | [-1.02, -0.03] |
| <sup>a</sup> Reference is Pittsburgh Site |  |  |  |  |  |  |
| <sup>b</sup> Reference is Men |  |  |  |  |  |  |
| <sup>c</sup> Reference is <HS/HS |  |  |  |  |  |  |
| <sup>d</sup> Reference is Never Smoker |  |  |  |  |  |  |

**Table 2A.** Linear regression results for women examining the relationship between childhood adverse life events and standardized ATP_Max_.

|  | **Model 1** |  | **Model 2** |  | **Model 3** |  |
| --- | --- | --- | --- | --- | --- | --- |
|  | **β** | **(95% CI)** | **β** | **(95% CI)** | **β** | **(95% CI)** |
| **Intercept** | 1.15 | [-0.23, 2.54] | 1.09 | [-0.36, 2.55] | 0.81 | [-0.88, 2.51] |
| **Childhood Adverse Life Events (cALEs)** | -0.04 | [-0.10, 0.02] | -0.04 | [-0.10, 0.02] | -0.05 | [-0.11, 0.01] |
| **Age (years)** | -0.02 | [-0.03, 0.00] | -0.02 | [-0.04, 0.00] | -0.01 | [-0.03, 0.01] |
| **Site<sup>a</sup>** | 0.12 | [-0.05, 0.30] | 0.15 | [-0.03, 0.33] | 0.13 | [-0.05, 0.31] |
| **Parental Education<sup>b</sup>** |  |  |  |  |  |  |
| Some college |  |  | -0.13 | [-0.38, 0.12] | -0.16 | [-0.41, 0.08] |
| College |  |  | 0.13 | [-0.14, 0.41] | 0.04 | [-0.24, 0.32] |
| Post College |  |  | 0.03 | [-0.25, 0.31] | 0.00 | [-0.29, 0.29] |
| **Body Mass Index (kg/m<sup>2</sup>)** |  |  |  |  | -0.03 | [-0.05, -0.01] |
| **CESD-10 Score (0-30 points)** |  |  |  |  | 0.03 | [-0.00, 0.05] |
| **Education<sup>b</sup>** |  |  |  |  |  |  |
| Some college |  |  |  |  | 0.11 | [-0.19, 0.41] |
| College |  |  |  |  | 0.24 | [-0.09, 0.57] |
| Post College |  |  |  |  | 0.03 | [-0.27, 0.33] |
| **Physical activity (cal/wk)** |  |  |  |  | 0.00 | [0.00, 0.00] |
| **Multimorbidity index (0-12)** |  |  |  |  | -0.03 | [-0.15, 0.09] |
| **Smoking Status<sup>c</sup>** |  |  |  |  |  |  |
| Former |  |  |  |  | -0.04 | [-0.23, 0.14] |
| Current |  |  |  |  | -0.45 | [-1.05, 0.15] |
| <sup>a</sup> Reference is Pittsburgh Site |  |  |  |  |  |  |
| <sup>b</sup> Reference is <HS/HS |  |  |  |  |  |  |
| <sup>c</sup> Reference is Never Smoker |  |  |  |  |  |  |

### Results for men

Among men, each additional adverse life event experienced before the age of 18 was borderline associated with -0.11SD lower ATP_max_ (95% CI= -0.21, -0.00). There was no change in the effect estimate which remained statistically significant with the inclusion of parental education. The effect estimate was slightly magnified when including participant education, smoking, physical activity, medication use, BMI and depressive symptoms (Model 3), such that each additional adverse life event was associated with -0.12 SD lower ATP_max_ (95% CI= -0.23, - 0.01) (Table 2B).

**Table 2B.**
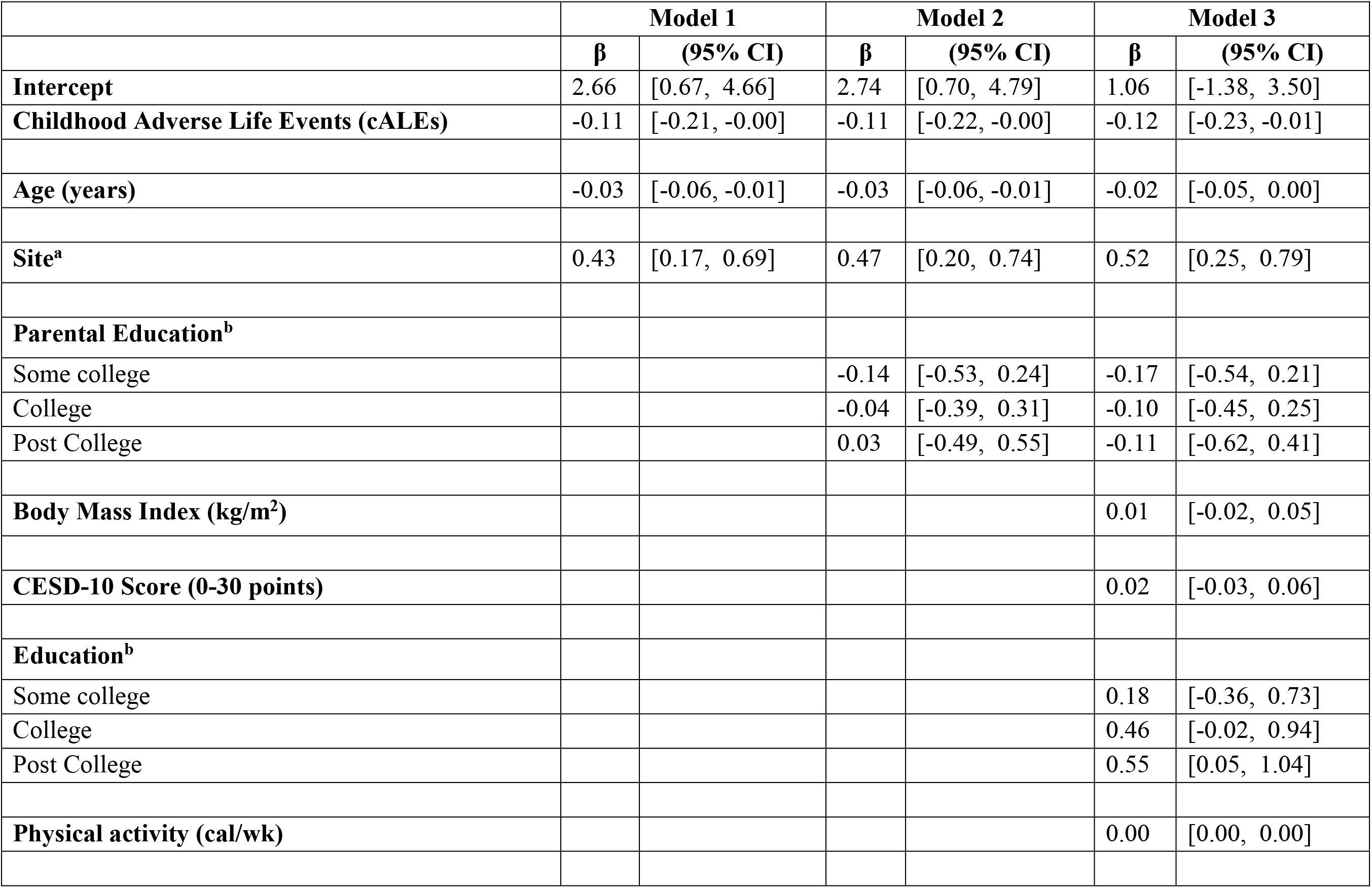
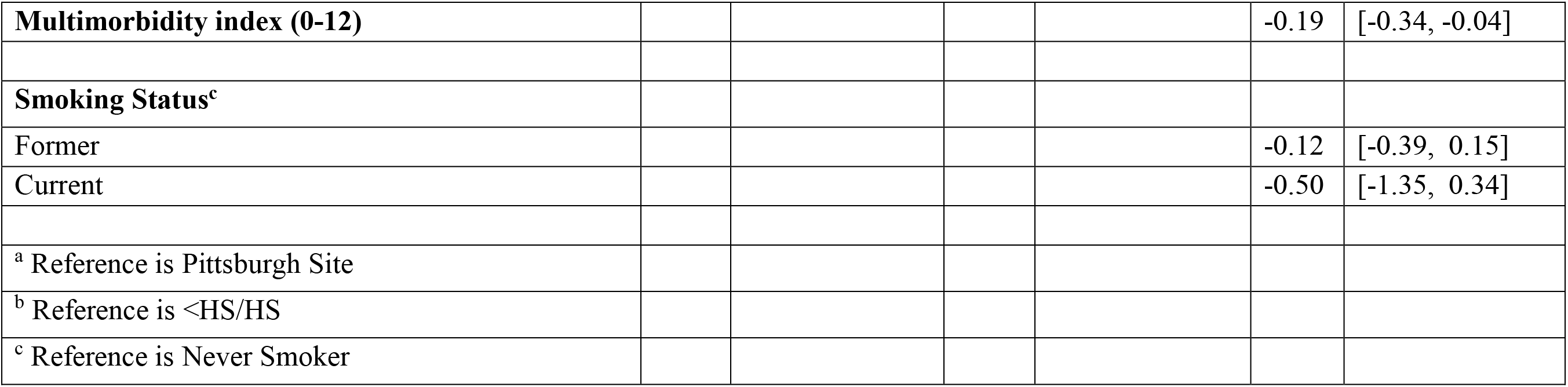
Linear regression results for men examining the relationship between childhood adverse life events and standardized ATP_Max_.

Associations were not statistically significant for Max OXPHOS measure for either women or men and are reported in Supplementary Table 1.

## DISCUSSION

In this well-characterized sample of older adults with gold-standard measures of mitochondrial bioenergetics, we investigated whether adverse life events experienced before the age of 18 were associated with reduced skeletal muscle mitochondrial function in later life. We found that in the overall sample, each additional adverse life event was associated with modestly lower ATP_max_, an in-vivo measure of maximal capacity for ATP production. In gender stratified models, significant associations between cALEs and ATP_max_ were observed in men but not in women. In contrast, no association was observed between childhood adverse life events and the *ex vivo* maximum oxidative phosphorylation capacity measure in either men or in women.

Few studies have explored whether stressful experiences in early life are associated with reduced mitochondrial function, recently recognized as a “hallmark of aging”,^31^ and implicated in the development of host of aging-related outcomes.^32–35^ Several studies have shown how stressful life experiences influence whole-body phenomenon such as allostatic load and inflammation,^36–38^ however the biologic processes by which these stressful social events experienced early in the life course may lead to impaired skeletal muscle mitochondria bioenergetics has received little attention, especially in the context of human cohort studies with both *ex vivo* and *in vivo* measures of bioenergetic function.

The results of our study offer preliminary evidence for the specific role skeletal muscle mitochondria may play in how adverse events become biologically embedded. While a small number of studies have examined how stressful experiences influence mitochondrial function, to date, much of this previous work has focused on alterations in content of mitochondrial DNA (mt-DNA). For example, adults who have experienced maltreatment in childhood or lost a parent before age 18 had significantly lower mt-DNA copy number compared to those with no adverse events in childhood.^14^ In close to 12,000 Chinese women with and without a history of major depression, childhood sexual abuse was shown to be associated with greater mt-DNA copy number in cells obtained from saliva samples.^39^ In a study of 290 adults, after controlling for perceived stress, resilience, depressive symptoms, and anxiety, the positive relationship between childhood adverse events and salivary mt-DNA copy number remained statistically significant.^10^ Taken together, given that the mt-DNA encodes proteins that influence cellular respiration and other bioenergetic properties, these results provide supporting evidence that core mitochondrial functions may be impaired in the face of social adversity, particularly those events experienced early in the life course.

We found that maximal ATP production in muscle was compromised among those who had experienced adverse life events in childhood, even after substantial adjustment. One possible explanation for this finding may lie in a what Picard and McEwen defined as “mitochondrial allostatic load” (MAL), a phenomenon in which mitochondria respond to social stressors.^11,40^ Specifically, MAL has been described as multifactorial, dynamic process by which mitochondria exhibit decreased respiratory enzymatic activity or mitochondrial membrane potential, which may lead to reduced energy production. This physiologic response is consistent with our finding that ATP_max_, a measure of maximal energy production, was lower among those who had experienced greater adverse events in childhood.

We investigated whether the relationship between cALEs and mitochondrial function differed by gender. While we did not find evidence of statistical interaction, we found in gender-stratified models, among men, each additional adverse life event in childhood was associated with lower ATP_max_. Although few studies have investigated whether there are gender differences in mitochondrial function in humans, a small body of evidence suggests that sex hormones may shape skeletal muscle metabolism, which may also influence the fiber type composition/size and capillary density.^41^ Therefore, it is possible that the inability to control for these confounding variables may in part explain why we observed different results by gender. An alternative reason for this difference may be driven by differential reporting. Prior research indicates that while women are more likely to suffer from post-traumatic stress disorder, men are more likely to report exposure to traumatic events.^42–44^

One unexpected finding was the lack of an observed association between the Max OXPHOS measure and cALEs. There are several potential explanations for this finding. First, while experiencing social stress may be related to a reduced capacity to generate ATP,^45,46^ this may occur without impairment to the electron transport system, as assessed by respirometry. Specifically, ATP_max_ is an integrated measure that calculates ATP production while the ex vivo Max OXPHOS measure reports oxygen consumption as a proxy for ATP production. If there is reduced coupling of oxygen consumption to ATP production (lower ratio of phosphate to oxygen) it could manifest in lower ATP_max_ without influencing the ex vivo Max OXPHOS measure.^48^ Changes in intracellular calcium cycling is another factor beyond respiratory chain capacity that could limit ATP_max_ but would not influence assays of mitochondrial respiration. Thus, the respiration assay used to determine Max OXPHOS utilizes oxygen and substrate concentrations that are standardized and saturating where there is no calcium cycling, which represents a fundamentally different assay compared to the ATP_max_ assay.^50^ Lastly, it is possible that measurement error may have been introduced between the two protocols which could explain the observed differences.

This study had several strengths. First, we utilized a well characterized sample of older adults that contained both in-vivo and ex vivo measures of mitochondrial function across a large number of participants. To the best of our knowledge, no studies have obtained gold-standard measures of mitochondrial function in a sample of this size. Second, we were also able to utilize life course social exposure data (i.e., parental education, educational attainment, adverse events experienced in childhood). The merging of gold-standard mitochondrial bioenergetic measures with highly valid and well-regarded measures of the social environment often do not exist within the same participants. Therefore, by leveraging this unique intersection of data, we were able to link adverse life events experienced in childhood, a well-established social exposure,^2,51,52^ to biologically specific measures of aging. Lastly, this study represents an important step forward in our understanding of how mitochondrial function, and specifically ATP production, measured in a tissue with high relevance for cardiometabolic health and physical function, may be implicated in compromised health among older adults later in the life course. Our study results provide initial mechanistic evidence for how early social stress may influence physical functioning and disability in later life,^5,53,54^ although future studies are needed to corroborate these findings.

Despite these strengths, this study has several limitations. First, we acknowledge the cross-sectional nature of these data which limits our ability to disentangle the temporal ordering of mediators on the causal pathway. In future studies, we hope to leverage longitudinal data, a recent addition to the SOMMA study, to parse the contribution of intervening factors that link childhood stress to adult health. Second, an additional limitation of the current study is the retrospective assessment of our primary exposure, childhood adverse life events, and the length of time between the cALEs and the assessment of mitochondrial function. Participants’ memory may be subject to recall bias if events that occurred earlier in life were more difficult to recall. However, previous research suggests individuals recall the timing of past traumatic events with reasonable accuracy.^53,55^ Third, we note that we treated each adverse event with equal weight and do not know the precise timing in which these events occurred (i.e., age 8 vs 18). Thus, we are unable to fully quantify the magnitude or severity of these adverse events across participants. Future work should pinpoint the duration, timing, and severity of the event experienced. Forth, our sample was predominantly non-Hispanic white and highly educated, limiting the generalizability of our findings. Fifth, we acknowledge the potential for residual confounding as we do not have data on the period between childhood and older age to control for indirect pathways by which adverse life events may have influenced mitochondrial function. Sixth, it is possible our estimates reflect overadjustment since several of the covariates included in our models, while potential confounders, could plausibly be hypothesized as mediators as well (i.e., education).^30^ However, even with the inability to assess the contribution of other variables on the causal pathway and the potential for over-adjustment due to controlling for mediators, we nonetheless observed an association between cALEs and ATP_max_. Our findings serve as a springboard for future studies to investigate how social stress experienced early in the life course influence mitochondrial function in later life, ideally in larger samples sample sizes.

## Conclusion

There is growing interest in understanding the social determinants of mitochondrial function among older adults. While few studies have investigated the role of early life social stress and mitochondrial function, almost no prior work has evaluated how adverse life events experienced in childhood influences mitochondrial function in a large sample of older adults. Therefore, this study is an important contribution to the literature because it assesses the relationship between cALEs and mitochondrial function in a tissue with high relevance for cardiometabolic health and physical function in among older adults. Our results suggest that adverse social stress experienced during childhood may be implicated in ATP production, particularly among men. Future work should examine these findings in the context of other relevant biologic pathways, including, but not limited to, inflammation, cell senescence and mt-DNA mutations and to what extent other mediating factors may also play a role in shaping energy production among older adults across the life course.

## Data availability

All data produced in the present study are available upon reasonable request to the authors

## SUPPLEMENTARY FILE

### Adapted from Adverse Childhood Experiences questionnaire

While you were growing up (under 18) did you experience any of the following?

1. Did someone in your close family ever use drugs or alcohol in a way that caused you to worry?
2. Did a parent or other adult in household ever swear at you, insult you, or put you down?
3. Did a parent or other adult in household ever slap, hit, beat, kick, or physically hurt you in any way?
4. Did you often feel that no one in your family loved you or thought you were important or special?
5. Did your parents or adults in your home ever slap, hit, beat, kick, or physically hurt each other?
6. Was either one of your parents absent from your life for a long period of time?

Question Responses:

1. Yes
2. No
3. Don’t know
4. Prefer not to answer

---

**Supplementary Figure 1.**
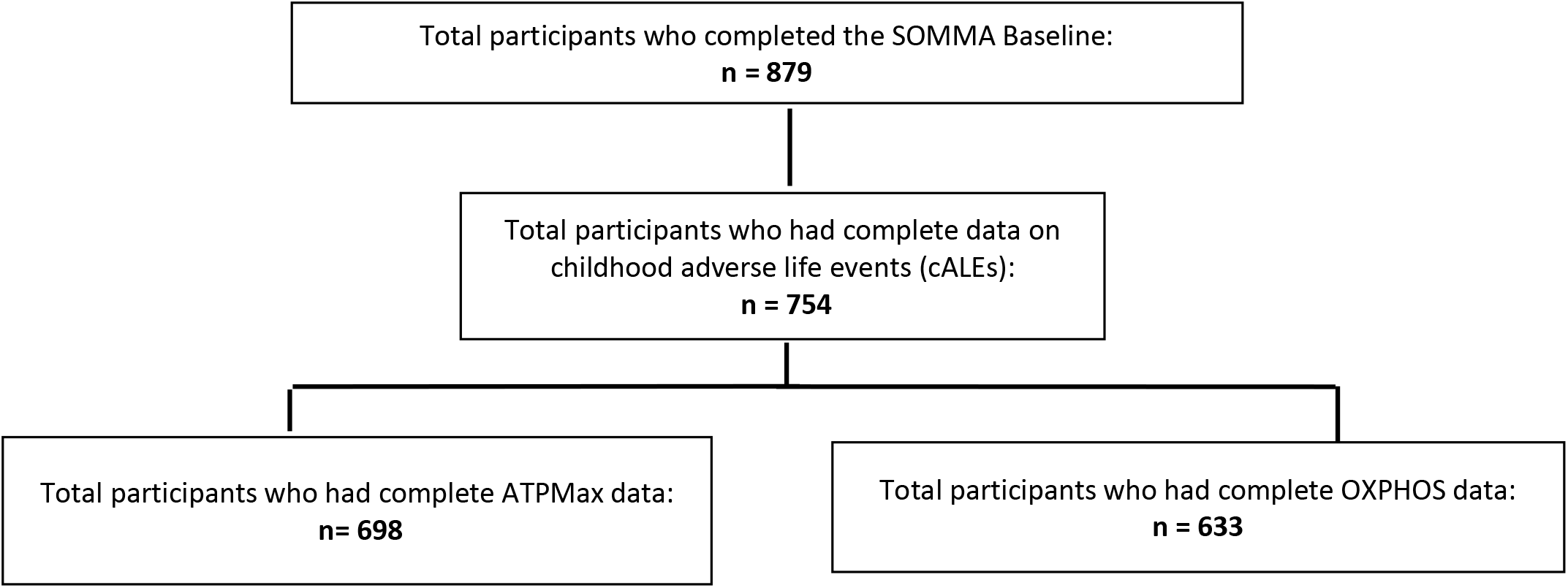
Sample flow among SOMMA participants with complete exposure and outcome data.

**Supplementary Table 1.** Overall linear regression results examining the relationship between childhood adverse life events and standardized Max OXPHOS.

|  | **Model 1** |  | **Model 2** |  | **Model 3** |  |
| --- | --- | --- | --- | --- | --- | --- |
|  | **β** | **(95% CI)** | **β** | **(95% CI)** | **β** | **(95% CI)** |
| **Intercept** | 2.12 | [0.95, 3.29] | 2.20 | [1.00, 3.40] | 2.55 | [1.11, 3.99] |
| **Childhood Adverse Life Events (cALEs)** | -0.05 | [-0.14, 0.04] | -0.04 | [-0.09, 0.02] | -0.03 | [-0.09, 0.02] |
| **Age (years)** | -0.02 | [-0.04, -0.01] | -0.02 | [-0.04, -0.01] | -0.02 | [-0.04, -0.01] |
| **Gender<sup>a</sup>** | -0.55 | [-0.74, -0.36] | -0.53 | [-0.68, -0.37] | -0.50 | [-0.66, -0.34] |
| **Gender\*cALEs** | 0.02 | [-0.09, 0.12] |  |  |  |  |
| **Technician<sup>b</sup>** |  |  |  |  |  |  |
| 1 | -0.48 | [-0.70, -0.26] | -0.49 | [-0.72, -0.27] | -0.47 | [-0.70, -0.24] |
| 2 | 0.58 | [0.14, 1.01] | 0.60 | [0.16, 1.03] | 0.51 | [0.08, 0.94] |
| 3 | 0.23 | [0.05, 0.41] | 0.22 | [0.03, 0.40] | 0.13 | [-0.05, 0.31] |
| 4 | 0.15 | [-0.16, 0.45] | 0.22 | [-0.10, 0.53] | 0.28 | [-0.02, 0.59] |
| **Parental Education<sup>c</sup>** |  |  |  |  |  |  |
| Some college |  |  | 0.11 | [-0.10, 0.33] | 0.12 | [-0.09, 0.33] |
| College |  |  | 0.21 | [0.00, 0.42] | 0.20 | [-0.02, 0.41] |
| Post College |  |  | -0.14 | [-0.39, 0.11] | -0.19 | [-0.45, 0.07] |
| **Body Mass Index (kg/m<sup>2</sup>)** |  |  |  |  | -0.03 | [-0.04, -0.01] |
| **CESD-10 Score (0-30 points)** |  |  |  |  | 0.00 | [-0.02, 0.02] |
| **Education<sup>c</sup>** |  |  |  |  |  |  |
| Some college |  |  |  |  | 0.09 | [-0.19, 0.37] |
| College |  |  |  |  | 0.08 | [-0.20, 0.35] |
| Post College |  |  |  |  | 0.13 | [-0.14, 0.40] |
| **Physical activity (cal/wk)** |  |  |  |  | 0.00 | [0.00, 0.00] |
| **Multimorbidity index (0-12)** |  |  |  |  | -0.12 | [-0.22, -0.03] |
| **Smoking Status<sup>d</sup>** |  |  |  |  |  |  |
| Former |  |  |  |  | 0.00 | [-0.15, 0.15] |
| Current |  |  |  |  | -0.33 | [-0.84, 0.18] |
| <sup>a</sup> Reference is Men |  |  |  |  |  |  |
| <sup>b</sup> Reference is Technician 5 |  |  |  |  |  |  |
| <sup>c</sup> Reference is <HS/HS |  |  |  |  |  |  |
| <sup>d</sup> Reference is Never Smoker |  |  |  |  |  |  |

**Supplementary Table 1A.** Linear regression results examining the relationship between childhood adverse life events and standardized Max OXPHOS among women.

|  | **Model 1** |  | **Model 2** |  | **Model 3** |  |
| --- | --- | --- | --- | --- | --- | --- |
|  | **β** | **(95% CI)** | **β** | **(95% CI)** | **β** | **(95% CI)** |
| **Intercept** | 0.59 | [-0.85, 2.03] | 0.48 | [-1.01, 1.98] | 0.97 | [-0.84, 2.78] |
| **Childhood Adverse Life Events (cALEs)** | -0.03 | [-0.09, 0.03] | -0.01 | [-0.08, 0.05] | -0.01 | [-0.08, 0.05] |
| **Age (years)** | -0.01 | [-0.03, 0.01] | -0.01 | [-0.03, 0.01] | -0.01 | [-0.03, 0.01] |
| **Technician<sup>a</sup>** |  |  |  |  |  |  |
| 1 | -0.50 | [-0.76, -0.25] | -0.49 | [-0.76, -0.22] | -0.44 | [-0.72, -0.16] |
| 2 | 0.38 | [-0.14, 0.90] | 0.44 | [-0.08, 0.95] | 0.36 | [-0.16, 0.87] |
| 3 | 0.33 | [0.12, 0.55] | 0.32 | [0.09, 0.54] | 0.25 | [0.03, 0.47] |
| 4 | 0.22 | [-0.15, 0.59] | 0.37 | [-0.02, 0.76] | 0.40 | [0.01, 0.78] |
| **Parental Education<sup>b</sup>** |  |  |  |  |  |  |
| Some college |  |  | 0.03 | [-0.23, 0.29] | 0.05 | [-0.20, 0.31] |
| College |  |  | 0.32 | [0.05, 0.59] | 0.32 | [0.04, 0.61] |
| Post College |  |  | -0.08 | [-0.36, 0.20] | -0.04 | [-0.35, 0.26] |
| **Body Mass Index (kg/m<sup>2</sup>)** |  |  |  |  | -0.02 | [-0.04, -0.00] |
| **CESD-10 Score (0-30 points)** |  |  |  |  | 0.01 | [-0.02, 0.03] |
| **Education<sup>b</sup>** |  |  |  |  |  |  |
| Some college |  |  |  |  | -0.10 | [-0.42, 0.22] |
| College |  |  |  |  | -0.10 | [-0.44, 0.24] |
| Post College |  |  |  |  | -0.15 | [-0.47, 0.17] |
| **Physical activity (cal/wk)** |  |  |  |  | 0.00 | [0.00, 0.00] |
| **Multimorbidity index (0-12)** |  |  |  |  | -0.08 | [-0.21, 0.04] |
| **Smoking Status<sup>c</sup>** |  |  |  |  |  |  |
| Former |  |  |  |  | 0.02 | [-0.17, 0.21] |
| Current |  |  |  |  | -0.23 | [-0.82, 0.37] |
| <sup>a</sup> Reference is Technician 5 |  |  |  |  |  |  |
| <sup>b</sup> Reference is <HS/HS |  |  |  |  |  |  |
| <sup>c</sup> Reference is Never Smoker |  |  |  |  |  |  |

**Supplementary Table 1B.**
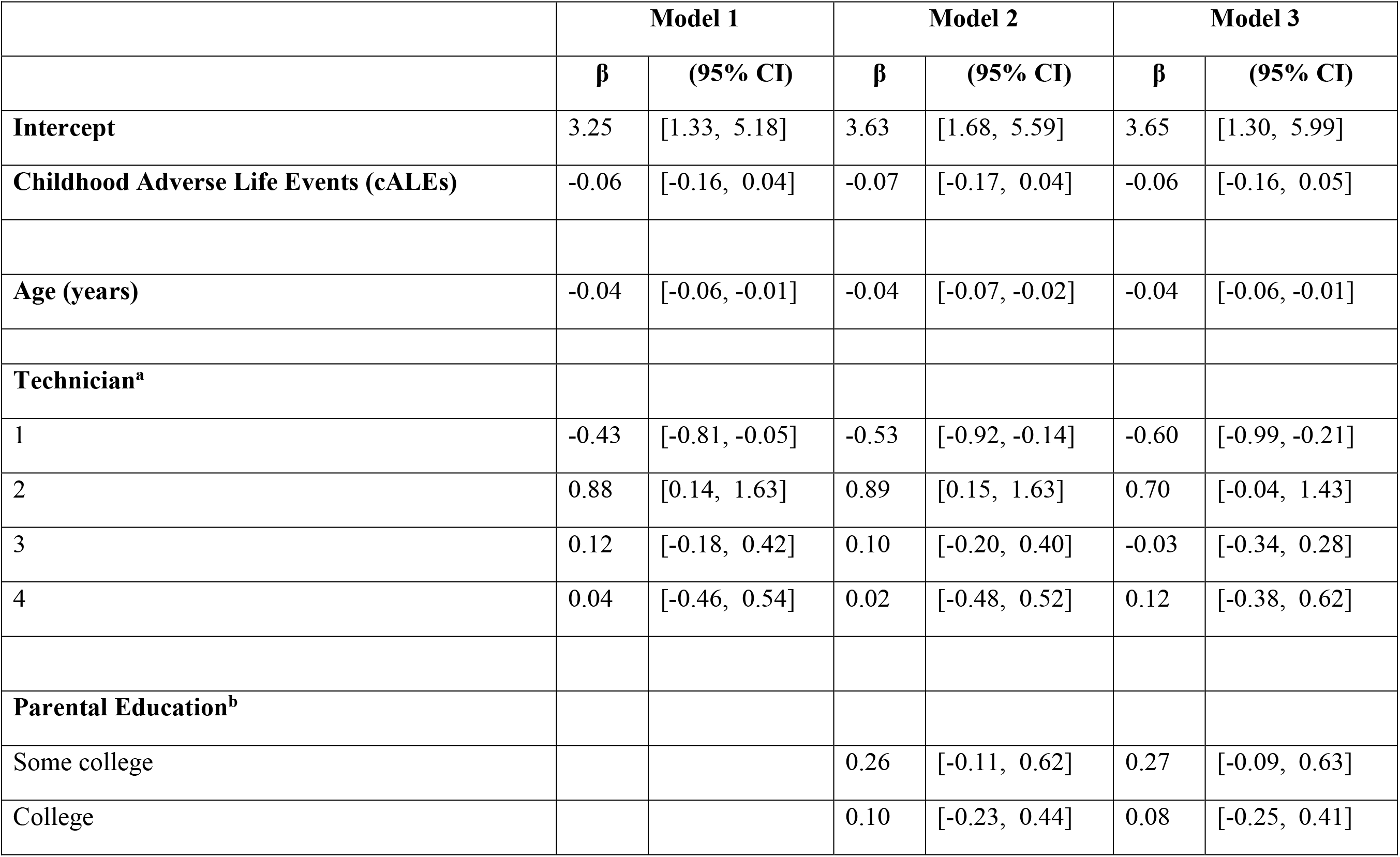
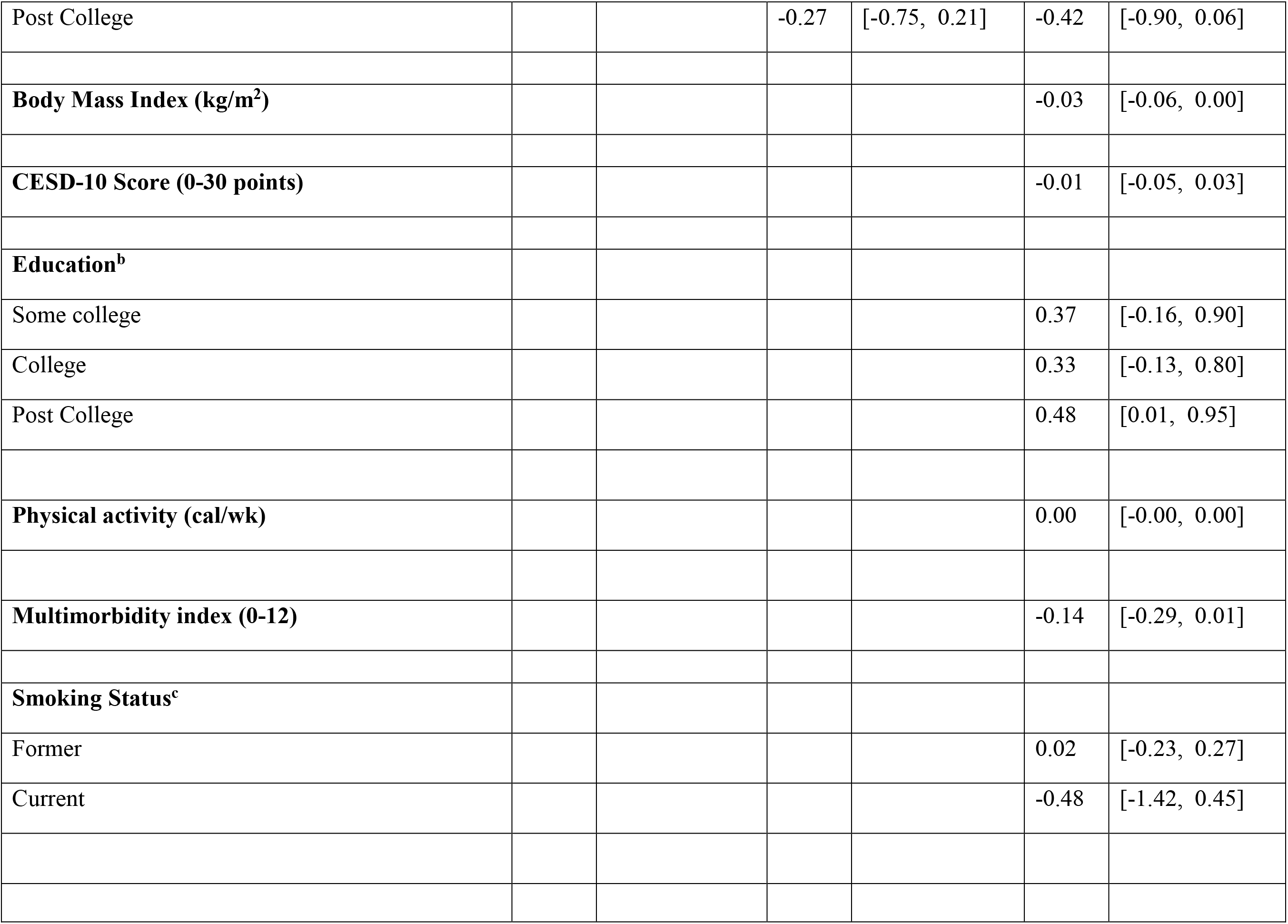
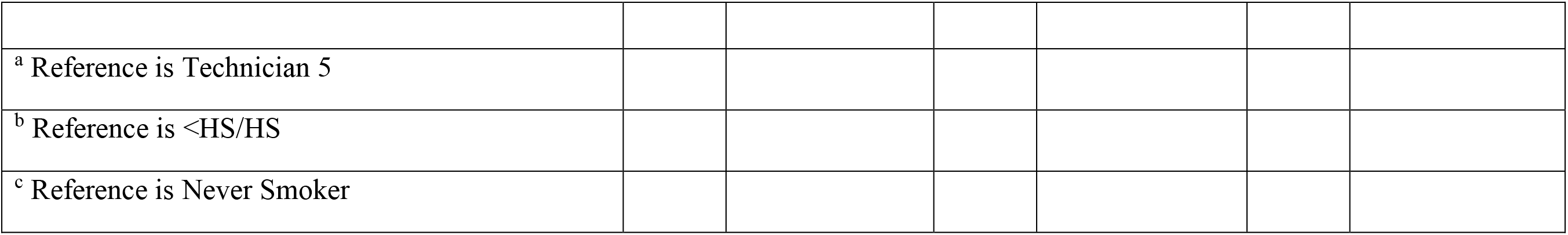
Linear regression results examining the relationship between childhood adverse life events and standardized Max OXPHOS among men.

## References

1. Kuh D, Ben-Shlomo Y, Lynch J, Hallqvist J, Power C. Life course epidemiology. J Epidemiol Community Health. 2003;57(10):778–783. doi:10.1136/JECH.57.10.778

2. Goodwin RD, Stein MB. Association between childhood trauma and physical disorders among adults in the United States. Psychol Med. 2004;34(3):509–520. doi:10.1017/S003329170300134X

3. Hayward MD, Gorman BK, Gorman BK, Hayward MD. The Long Arm of Childhood: The Influence of Early-Life Social Conditions on Men’s Mortality. Demography. 2004;41(1):87–107. doi:10.1353/dem.2004.0005

4. Ridout KK, Khan M, Ridout SJ. Adverse Childhood Experiences Run Deep: Toxic Early Life Stress, Telomeres, and Mitochondrial DNA Copy Number, the Biological Markers of Cumulative Stress. BioEssays. 2018;40(9):1800077. doi:10.1002/bies.201800077

5. Duchowny KA, Hicken MT, Cawthon PM, Glymour MM, Clarke P. Life course trauma and muscle weakness in older adults by gender and race/ethnicity: Results from the U.S. health and Retirement Study. SSM - Popul Health. 2020;11:100587. doi:10.1016/j.ssmph.2020.100587

6. Martínez J, Marmisolle I, Tarallo D, Quijano C. Mitochondrial Bioenergetics and Dynamics in Secretion Processes. Front Endocrinol. 2020;11. Accessed February 7, 2023. https://www.frontiersin.org/articles/10.3389/fendo.2020.00319

7. Picard M, McEwen BS. Psychological Stress and Mitochondria: A Systematic Review. Psychosom Med. 2018;80(2):141–153. doi:10.1097/PSY.0000000000000545

8. Picard M, McManus MJ, Gray JD, et al. Mitochondrial functions modulate neuroendocrine, metabolic, inflammatory, and transcriptional responses to acute psychological stress. Proc Natl Acad Sci. 2015;112(48):E6614–E6623. doi:10.1073/pnas.1515733112

9. Zitkovsky EK, Daniels TE, Tyrka AR. Mitochondria and early-life adversity. Mitochondrion. 2021;57:213–221. doi:10.1016/j.mito.2021.01.005

10. Tyrka AR, Parade SH, Price LH, et al. Alterations of Mitochondrial DNA Copy Number and Telomere Length With Early Adversity and Psychopathology. Biol Psychiatry. 2016;79(2):78–86. doi:10.1016/j.biopsych.2014.12.025

11. Picard M, Juster RP, McEwen BS. Mitochondrial allostatic load puts the “gluc” back in glucocorticoids. Nat Rev Endocrinol. 2014;10(5):303–310. doi:10.1038/nrendo.2014.22

12. Picard M, McEwen BS. Psychological Stress and Mitochondria: A Conceptual Framework. Psychosom Med. 2018;80(2):126–140. doi:10.1097/PSY.0000000000000544

13. Picard M, Prather AA, Puterman E, et al. A Mitochondrial Health Index Sensitive to Mood and Caregiving Stress. Biol Psychiatry. 2018;84(1):9–17. doi:10.1016/j.biopsych.2018.01.012

14. Boeck C, Koenig AM, Schury K, et al. Inflammation in adult women with a history of child maltreatment: The involvement of mitochondrial alterations and oxidative stress. Mitochondrion. 2016;30:197–207. doi:10.1016/j.mito.2016.08.006

15. Gumpp AM, Boeck C, Behnke A, et al. Childhood maltreatment is associated with changes in mitochondrial bioenergetics in maternal, but not in neonatal immune cells. Proc Natl Acad Sci. 2020;117(40):24778–24784. doi:10.1073/pnas.2005885117

16. Tyrka AR, Parade SH, Price LH, et al. Alterations of Mitochondrial DNA Copy Number and Telomere Length with Early Adversity and Psychopathology. Biol Psychiatry. 2016;79(2):78–86. doi:10.1016/j.biopsych.2014.12.025

17. Cummings SR, Newman AB, Coen PM, et al. The Study of Muscle, Mobility and Aging (SOMMA). A Unique Cohort Study about the Cellular Biology of Aging and Age-related Loss of Mobility. J Gerontol A Biol Sci Med Sci. Published online February 9, 2023:glad052. doi:10.1093/gerona/glad052

18. Dube SR, Anda RF, Felitti VJ, Chapman DP, Williamson DF, Giles WH. Childhood Abuse, Household Dysfunction, and the Risk of Attempted Suicide Throughout the Life SpanFindings From the Adverse Childhood Experiences Study. JAMA. 2001;286(24):3089–3096. doi:10.1001/jama.286.24.3089

19. Mau T, Lui LY, Distefano G, et al. Mitochondrial energetics in skeletal muscle are associated with leg power and cardiorespiratory fitness in the Study of Muscle, Mobility, and Aging (SOMMA). J Gerontol A Biol Sci Med Sci. Published online December 3, 2022:glac238. doi:10.1093/gerona/glac238

20. Jubrias SA, Crowther GJ, Shankland EG, Gronka RK, Conley KE. Acidosis inhibits oxidative phosphorylation in contracting human skeletal muscle in vivo. J Physiol. 2003;553(Pt 2):589–599. doi:10.1113/jphysiol.2003.045872

21. Blei ML, Conley KE, Kushmerick MJ. Separate measures of ATP utilization and recovery in human skeletal muscle. J Physiol. 1993;465:203–222. doi:10.1113/jphysiol.1993.sp019673

22. Amara CE, Marcinek DJ, Shankland EG, Schenkman KA, Arakaki LSL, Conley KE. Mitochondrial function in vivo: spectroscopy provides window on cellular energetics. Methods San Diego Calif. 2008;46(4):312–318. doi:10.1016/j.ymeth.2008.10.001

23. Currie CE, Elton RA, Todd J, Platt S. Indicators of socioeconomic status for adolescents: the WHO Health Behaviour in School-aged Children Survey. Health Educ Res. 1997;12(3):385–397. doi:10.1093/her/12.3.385

24. Aarø LE, Flisher AJ, Kaaya S, Onya H, Namisi FS, Wubs A. Parental education as an indicator of socioeconomic status: improving quality of data by requiring consistency across measurement occasions. Scand J Public Health. 2009;37(Suppl. 2):16–27.

25. Lewinsohn PM, Seeley JR, Roberts RE, Allen NB. Center for Epidemiologic Studies Depression Scale (CES-D) as a screening instrument for depression among community-residing older adults. Psychol Aging. 1997;12(2):277–287. doi:10.1037//0882-7974.12.2.277

26. Stewart AL, Mills KM, King AC, Haskell WL, Gillis D, Ritter PL. CHAMPS physical activity questionnaire for older adults: outcomes for interventions. Med Sci Sports Exerc. 2001;33(7):1126–1141. doi:10.1097/00005768-200107000-00010

27. Vassilaki M, Aakre JA, Cha RH, et al. Multimorbidity and Risk of Mild Cognitive Impairment. J Am Geriatr Soc. 2015;63(9):1783–1790. doi:10.1111/jgs.13612

28. Silaidos C, Pilatus U, Grewal R, et al. Sex-associated differences in mitochondrial function in human peripheral blood mononuclear cells (PBMCs) and brain. Biol Sex Differ. 2018;9(1):34. doi:10.1186/s13293-018-0193-7

29. Miotto PM, McGlory C, Holloway TM, Phillips SM, Holloway GP. Sex differences in mitochondrial respiratory function in human skeletal muscle. Am J Physiol Regul Integr Comp Physiol. 2018;314(6):R909–R915. doi:10.1152/ajpregu.00025.2018

30. Schisterman EF, Cole SR, Platt RW. Overadjustment bias and unnecessary adjustment in epidemiologic studies. Epidemiol Camb Mass. 2009;20(4):488–495. doi:10.1097/EDE.0b013e3181a819a1

31. López-Otín C, Blasco MA, Partridge L, Serrano M, Kroemer G. The Hallmarks of Aging. Cell. 2013;153(6):1194–1217. doi:10.1016/j.cell.2013.05.039

32. López-Otín C, Blasco MA, Partridge L, Serrano M, Kroemer G. The Hallmarks of Aging. Cell. 2013;153(6):1194–1217. doi:10.1016/j.cell.2013.05.039

33. Chan DC. Mitochondria: Dynamic Organelles in Disease, Aging, and Development. Cell. 2006;125(7):1241–1252. doi:10.1016/J.CELL.2006.06.010

34. Santanasto AJ, Coen PM, Glynn NW, et al. The relationship between mitochondrial function and walking performance in older adults with a wide range of physical function. Exp Gerontol. 2016;81:1–7. doi:10.1016/j.exger.2016.04.002

35. Parry HA, Roberts MD, Kavazis AN. Human Skeletal Muscle Mitochondrial Adaptations Following Resistance Exercise Training. Int J Sports Med. 2020;41(6):349–359. doi:10.1055/a-1121-7851

36. Clark MS, Bond MJ, Hecker JR. Environmental stress, psychological stress and allostatic load. Psychol Health Med. 2007;12(1):18–30. doi:10.1080/13548500500429338

37. Juster RP, McEwen BS, Lupien SJ. Allostatic load biomarkers of chronic stress and impact on health and cognition. Neurosci Biobehav Rev. 2010;35(1):2–16. doi:10.1016/j.neubiorev.2009.10.002

38. Cuevas AG, Ong AD, Carvalho K, et al. Discrimination and systemic inflammation: A critical review and synthesis. Brain Behav Immun. 2020;89:465–479. doi:10.1016/j.bbi.2020.07.017

39. Cai N, Chang S, Li Y, et al. Molecular signatures of major depression. Curr Biol CB. 2015;25(9):1146–1156. doi:10.1016/j.cub.2015.03.008

40. Picard M, McEwen BS. Psychological Stress and Mitochondria: A Conceptual Framework. Psychosom Med. 2018;80(2):126–140. doi:10.1097/PSY.0000000000000544

41. Lundsgaard AM, Kiens B. Gender Differences in Skeletal Muscle Substrate Metabolism – Molecular Mechanisms and Insulin Sensitivity. Front Endocrinol. 2014;5:195. doi:10.3389/fendo.2014.00195

42. Tolin DF, Foa EB. Sex differences in trauma and posttraumatic stress disorder: A quantitative review of 25 years of research. Psychol Trauma Theory Res Pract Policy. 2008;S(1):37–85. doi:10.1037/1942-9681.S.1.37

43. Kessler RC, Mickelson KD, Williams DR. The prevalence, distribution, and mental health correlates of perceived discrimination in the United States. J Health Soc Behav. 1999;40(3):208–230. doi:10.2307/2676349

44. Christiansen DM, Hansen M. Accounting for sex differences in PTSD: A multi-variable mediation model. Eur J Psychotraumatology. 2015;6:10.3402/ejpt.v6.26068. doi:10.3402/ejpt.v6.26068

45. Picard M, McEwen BS, Epel ES, Sandi C. An energetic view of stress: Focus on mitochondria. Front Neuroendocrinol. 2018;49:72–85. doi:10.1016/j.yfrne.2018.01.001

46. Hollis F, van der Kooij MA, Zanoletti O, Lozano L, Cantó C, Sandi C. Mitochondrial function in the brain links anxiety with social subordination. Proc Natl Acad Sci. 2015;112(50):15486–15491. doi:10.1073/pnas.1512653112

47. Djafarzadeh S, Jakob SM. High-resolution Respirometry to Assess Mitochondrial Function in Permeabilized and Intact Cells. J Vis Exp JoVE. 2017;(120):54985. doi:10.3791/54985

48. Layec G, Gifford JR, Trinity JD, et al. Accuracy and precision of quantitative 31P-MRS measurements of human skeletal muscle mitochondrial function. Am J Physiol - Endocrinol Metab. 2016;311(2):E358–E366. doi:10.1152/ajpendo.00028.2016

49. Tsampasian V, Cameron D, Sobhan R, Bazoukis G, Vassiliou VS. Phosphorus Magnetic Resonance Spectroscopy (31P MRS) and Cardiovascular Disease: The Importance of Energy. Medicina (Mex). 2023;59(1):174. doi:10.3390/medicina59010174

50. Cardinale DA, Gejl KD, Ørtenblad N, Ekblom B, Blomstrand E, Larsen FJ. Reliability of maximal mitochondrial oxidative phosphorylation in permeabilized fibers from the vastus lateralis employing high-resolution respirometry. Physiol Rep. 2018;6(4):e13611. doi:10.14814/phy2.13611

51. Baumeister D, Akhtar R, Ciufolini S, Pariante CM, Mondelli V. Childhood trauma and adulthood inflammation: a meta-analysis of peripheral C-reactive protein, interleukin-6 and tumour necrosis factor-α. Mol Psychiatry. 2016;21(5):642–649. doi:10.1038/mp.2015.67

52. Putnam KT, Harris WW, Putnam FW. Synergistic Childhood Adversities and Complex Adult Psychopathology. J Trauma Stress. 2013;26(4):435–442. doi:10.1002/jts.21833

53. Haas S. Trajectories of functional health: The “long arm” of childhood health and socioeconomic factors. Published online 2007. doi:10.1016/j.socscimed.2007.11.004

54. Birnie K, Cooper R, Martin RM, et al. Childhood Socioeconomic Position and Objectively Measured Physical Capability Levels in Adulthood: A Systematic Review and Meta-Analysis. Vitzthum V, ed. PLoS ONE. 2011;6(1):e15564. doi:10.1371/journal.pone.0015564

55. Krall EA, Valadian I, Dwyer JT, Gardner J. Recall of childhood illnesses. J Clin Epidemiol. 1988;41(11):1059–1064. doi:10.1016/0895-4356(88)90075-3

